# Regression for accurate and sensitive grading of mutations diagnostic of antibiotic resistance in *Mycobacterium tuberculosis*

**DOI:** 10.1101/2024.07.01.24309598

**Authors:** Sanjana G. Kulkarni, Sacha Laurent, Paolo Miotto, Timothy M. Walker, Leonid Chindelevitch, WHO sequencing network, Carl-Michael Nathanson, Nazir Ismail, Timothy C. Rodwell, Maha R. Farhat

## Abstract

Rapid genotype-based drug susceptibility testing for the *Mycobacterium tuberculosis* complex (MTBC) relies on a comprehensive knowledgebase of the genetic determinants of resistance. We built a catalog of resistance-associated mutations in MTBC using a novel regression-based approach and benchmarked it against the 2^nd^ edition of the World Health Organization mutation catalog. We trained multivariate logistic regression models on over 50,000 MTBC isolates to associate binary resistance phenotypes for 15 antitubercular drugs with variants extracted from candidate resistance genes. Regression detects 452/457 (99%) resistance-associated variants identified using the existing method (*a.k.a,* SOLO method) and grades 218 (29%) more total variants than SOLO. The regression-based catalog achieves higher sensitivity on average (+3.2 percentage points, pp) than SOLO with smaller average decreases in specificity (−1.0 pp) and positive predictive value (−1.8 pp). The regression pipeline also detects isoniazid resistance compensatory mutations in *ahpC* and variants linked to bedaquiline and aminoglycoside hypersusceptibility. These results inform the continued development of targeted next generation sequencing, whole genome sequencing, and other commercial molecular assays for diagnosing resistance in MTBC. In addition to grading genetic variants by their associations with phenotype, regression models could potentially provide an accurate and scalable method of predicting antibiotic resistance from bacterial genetic profiles.

## Introduction

The current method used by the World Health Organization to build catalogs of mutations associated with antibiotic resistance focuses on univariate association between solitary mutations and binary resistance phenotype (resistant vs. susceptible), excluding isolates with multiple possibly causative resistance mutations. Additional confidence grading rules are applied to the output of this association analysis and integrated with independent data from the literature (*e.g.* allelic exchange data) to generate the final grading of mutations into 5 categories: Group 1) Associated with resistance, Group 2) Associated with Resistance - Interim, Group 3) Uncertain significance, Group 4) Not associated with Resistance - Interim, and Group 5) Not associated with resistance.

This *univariate association* method^1,2^ is based upon the assumption that most resistant strains have just one non-synonymous, non-lineage defining mutation in candidate genes whereas most susceptible strains have none^2^. Evidence implicating a mutation with resistance is derived solely from isolates in which it is the only mutation occurring in a drug-resistant isolate (after exclusion of a pre-specified list of neutral variants). This approach considers the mutation independently of potential additive effects with other mutations. Mutations that do not fit those criteria are graded using additional grading rules *(i.e.* predicted effect on protein function, proximity to the drug’s active site) or evidence from the literature. If none of the above is possible, then it is graded as uncertain^3,4^.

Different from univariate association, multivariable regression can perform phenotypic association of solitary or multiple co-occurring mutations, estimating the effect of each mutation on phenotype, conditional on the presence or absence of other mutations. Regression is used to generate polygenic risk scores for predicting human disease^5^. Similarly, given the known high heritability of antibiotic resistance in bacteria^6,7^, this approach can also be used to predict antibiotic resistance from genotypic data. Both binary and semi-continuous outcomes (*e.g.* minimum inhibitory concentrations, or MICs, are typically measured at serial doubling dilutions) can be predicted. However, genetic association using regression requires an assumption that the relationship between mutation presence and the probability of phenotypic resistance follows a logistic function and that the effects of co-occurring mutations on resistance are additive. Strong co-linearity between mutations can bias the fit of a regression model and affect its attribution of effects between linked mutations and the phenotype. Further, accurate association using regression requires genotypic and phenotypic data from a large and diverse sample set.

Motivated by the potential advantages of regression for genotype to phenotype association and enabled by the WHO global collection of MTBC genomes (N = 52,567), we tested the accuracy and predictive performance of a multivariable penalized regression model compared with the SOLO method currently used in the World Health Organization (WHO) catalog of mutations in Mycobacterium tuberculosis complex and their association with drug resistance (“WHO mutation catalog”)^4^. This approach may also lower dependence on literature sources external to the data to grade mutations and improve scalability and future automation of catalog production.

## Methods

### A. Definitions, Abbreviations, and Curation of MTBC data

Regression models were trained using the same genotypic and phenotypic data as the 2^nd^ edition of the published WHO mutation catalog^4^. These phenotypic and genotypic data were curated from published datasets, consortium initiatives, and direct submissions in response to public calls for contributions by the WHO Global Tuberculosis Programme^8^. Quality control was previously performed for the 2^nd^ edition of the mutation catalog, with 52,567 isolates passing both phenotypic and genotypic data quality control^4^.

The pDST data were subset into two groups: the “WHO dataset” (N = 41,141) consists of higher confidence phenotypes tested using WHO-approved phenotypic testing methods, and the “ALL dataset” (N = 52,567) includes the entire WHO dataset as well as pDST results measured using phenotypic approaches not endorsed by the WHO (*e.g.* UKMYC 96-well plates)^9^.

Candidate resistance genes were assigned to two tiers based on published literature, including the first edition of the catalog^3^, and discussions among an international panel of expert advisors: Tier 1 is composed of genes and associated promoter regions deemed by the expert panel to contain resistance mutations with high probability and Tier 2 is composed of genes that may contain resistance mutations based on new or less established evidence^4^. We focused the regression on associating variants in Tier 1 genes only **(Supplementary Table 1)** because evidence from the 2^nd^ edition of the mutation catalog did not support the association of variants in Tier 2 genes with drug resistance^4^.

Variants not covered by at least 10 reads in the read alignment were labeled as missing calls^4^. For variants with at least 10 reads of support, a variant was considered present if it had a within-isolate allele frequency (AF) > 0.75 and absent if AF ≤ 0.25. We did not attempt to associate variants with intermediate allele frequencies with resistance in the regression models. Regression models cannot be trained on missing data, so for a given single-drug model, we excluded all isolates with variant calls **(Supplementary Table 2)** with an AF in the range (0.25, 0.75] and isolates with missing variant calls. After association, resistance prediction to assess sensitivity, specificity, and PPV did use isolates with intermediate allele frequencies as described in the results section. The genes *rrs* and *rrl* are known to have a high degree of homology across bacteria, and false low frequency variant calls are common due to contamination in the sequencing process. For this reason, the ribosome-targeting drugs have the largest numbers of excluded isolates and associated variants **(Supplementary Table 2)**.

### B. Regression Model Design

To maintain high statistical power, we built a series of nested models **(Supplementary Table 3)**. The “base” model for a given phenotypic group was fit on all non-silent variants with no pooling of LoF variants. Two additional models were fit: one in which LoF variants were pooled on a per-gene basis and another with silent variants in addition to unpooled non-silent variants. The expected effect size of silent variants was significantly smaller than those of non-silent mutations, so no model was trained only on silent variants. Definitions of silent variants and LoF mutations were the same as for the SOLO method **(Table 1)**. To pool LoFs, for a given gene, all frameshift, start_lost, stop_gained, and feature_ablation mutations were combined into a single “LoF” variant. The component mutations were then removed from the variant matrix so that a variant was not multiply considered during model fitting.

**Table 1.**
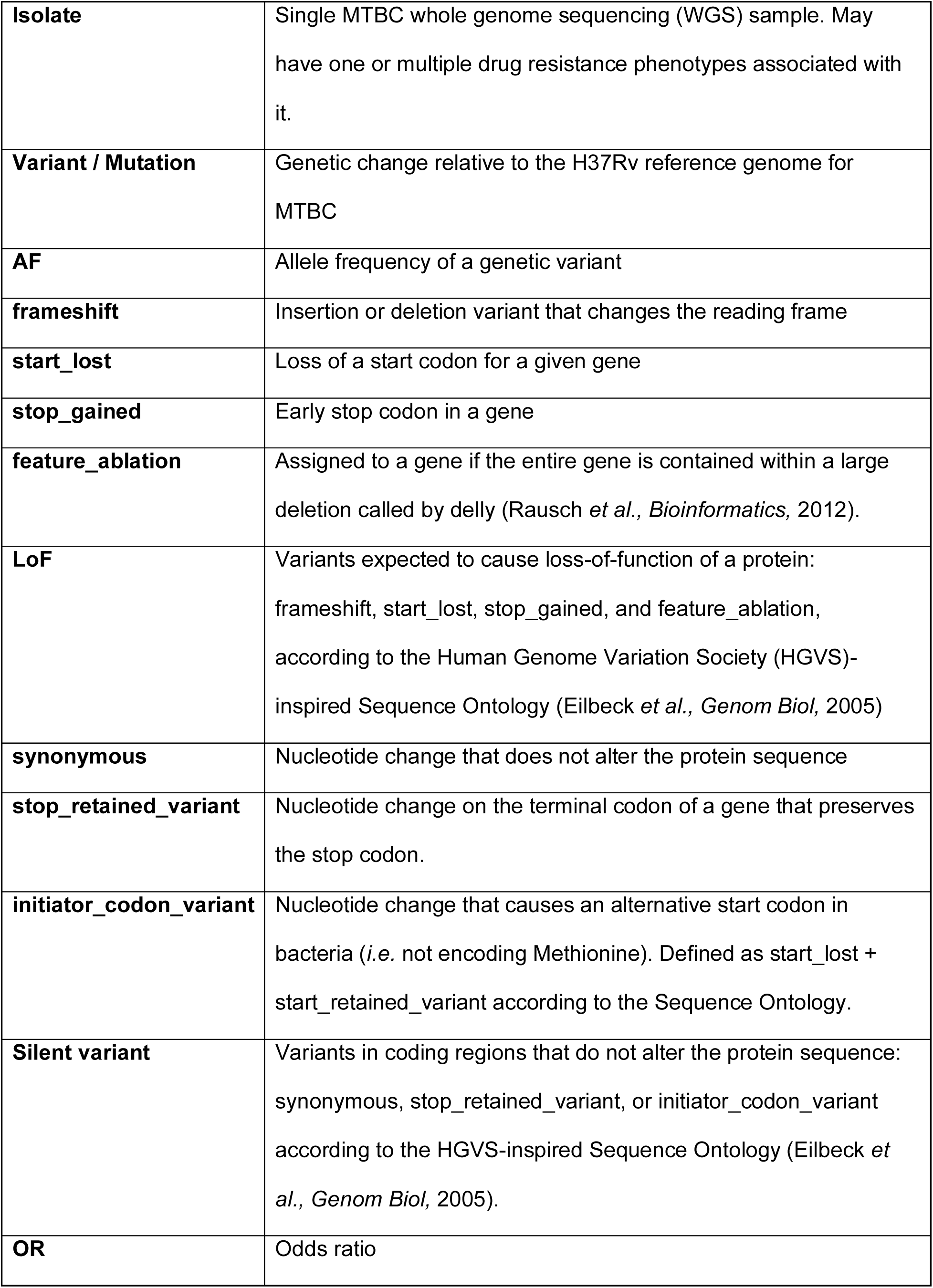
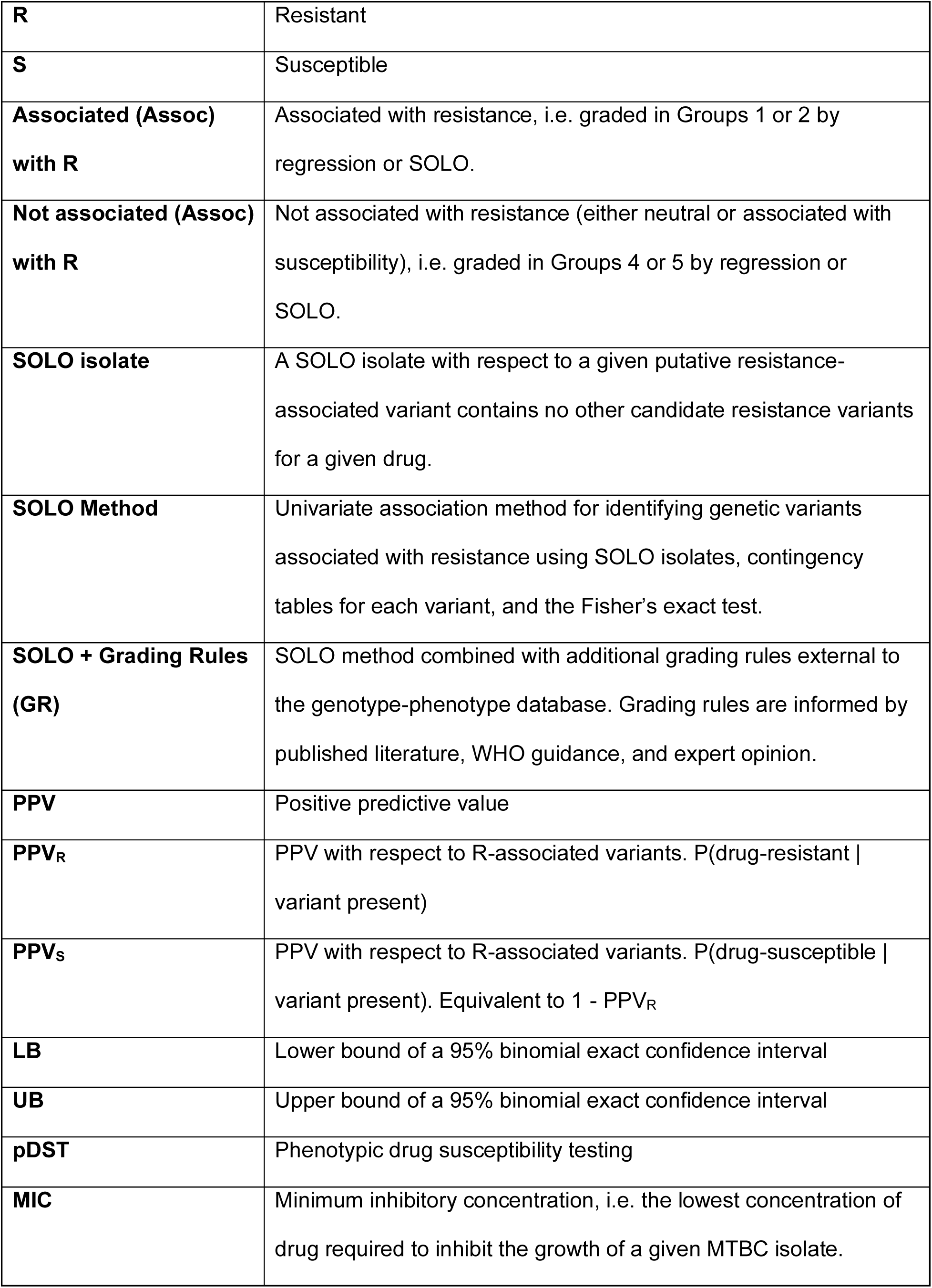
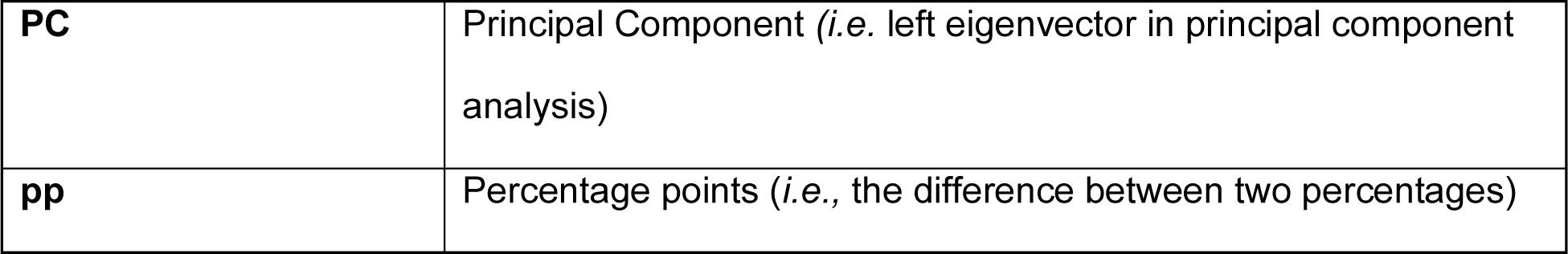
Definitions and abbreviations used throughout the manuscript. These are the same definitions and abbreviations as in the 2^nd^ edition of the MTBC resistance mutation catalog. Variant effect names are according to the Sequence Ontology^33^.

Non-silent non-indel variants were tested in all three models in **Supplementary Table 3,** but we used only the results from the unpooled model to grade them. The only results derived from the models with LoF pooling and silent variants were the pooled LoF and silent variants themselves, respectively. We separately fit models on the WHO and ALL datasets. Models were trained to regress single-drug binary pDST on genotype. The MIC models were not used for grading but were instead used to add supportive evidence for associations between genotype and pDST.

We trained single-drug regression models with L2 penalties due to expected multi-collinearity in the data, which is caused by clonality and population structure in bacteria. We performed a series of statistical tests to grade mutations into five categories. Because genes were selected for inclusion into the models based on the literature and known or putative mechanisms of action, all mutations in the genes in **Supplementary Table 1** were included. To select the strength of L2 regularization for each model, we performed five-fold cross-validation using all powers of ten from 10^-6^ to 10^6^ and selected the regularization parameter with the lowest binary cross-entropy, with class balancing. Coefficient significance was determined using a permutation test with 1000 re-shufflings of phenotype across the full dataset and the same regularization parameter determined on the original model **(Supplementary** Figure 1a-b).

### C. Principal Component Analysis

MTBC is a clonally evolving bacterium with strong population structure, which can lead to systematic false positives when estimating the effects of mutations on resistance that may not be sufficiently corrected for through L2 regularization^10,11^. Therefore, we additionally adjusted for population structure using principal components analysis (PCA) on a kinship matrix. 6,938 single nucleotide variants (SNVs) more than 50 base pairs away from any PE or PPE gene across the MTBC genome and with a variant allele frequency of at least 1% were included. We excluded previously identified homoplastic sites^12^, putative drug resistance regions^13^, and sites at which more than 1% of isolates have a variant occurring at a within-isolate read frequency <75%, leaving 6,190 SNV sites for the kinship matrix.

We performed PCA on the covariance matrix of these 6,190 sites. The resulting principal components are latent variables the first of which are expected to describe major axes of ancestral variation (*i.e.,* lineage) in the MTBC. We included the first 50 principal components, which explain 99.995 % of the total variance in the genotype, as covariates in every model. We ran fast-lineage-caller^14^ on the variant call format (VCF) files to obtain lineage designations **(Supplementary Table 4).** We considered an isolate’s lineage to be the most specific lineage that fast-lineage-caller identified. Eight isolates did not have a lineage by the Coll scheme^15^ but were classified as *Mycobacterium canettii* by the Lipworth scheme^16^. Several principal components are highly correlated with lineages and sublineages **(Supplementary** Figure 2). Many of the principal components **(Supplementary** Figure 2e-h) separate sublineages of L4, which is the most diverse of the MTBC lineages. 395/52,567 (0.75%) of isolates have more than one lineage assigned according to the Coll scheme^15^. These are excluded from **Supplementary** Figure 2 but are in all models and analyses.

### D. Likelihood Ratio Test

To further discriminate between the strengths of association of different mutations, we performed a likelihood ratio test (LRT). This test compares two nested models and determines if adding additional variants significantly improves a model’s goodness-of-fit. The LRT is more conservative than the permutation test as it more directly controls for collinearity in the mutations, *i.e.* for a mutation to demonstrate significance on the LRT it must improve the fit of the model when added to all other mutations in the model. This LRT statistic follows the chi-squared distribution **(Supplementary** Figure 1c).

### E. Test for Mutation Neutrality

We implemented a second permutation test that reverses the hypothesis test to determine if the evidence supports a lack of an association between a variant and drug resistance (*i.e.,* test if a mutation is neutral with an OR ∼1 on resistance). In the permutation test for mutation effect, if the proportion of permuted ORs that are more extreme than the estimated OR on the training dataset is less than α, then the variant has a significant OR on resistance **(Supplementary** Figure 1a). Conversely, for the test for mutation neutrality, if the proportion of permuted ORs that are less extreme (*i.e.,* closer to 1) than the estimated OR is less than α, then the variant is not associated with the drug resistance phenotype **(Supplementary** Figure 1b).

### F. Mutation Classifications

We used five grading groups as in the WHO catalog^4^, noting that these gradings were necessarily defined by different statistical criteria for regression. The flowchart in **Figure 1** details the grading for each variant in a single model *(i.e.,* single drug, single phenotypic group). The gradings assigned in **Figure 1** are interim, after which the gradings were integrated between the two phenotypic datasets to a final grading **(Supplementary Table 5).**

**Figure 1.**
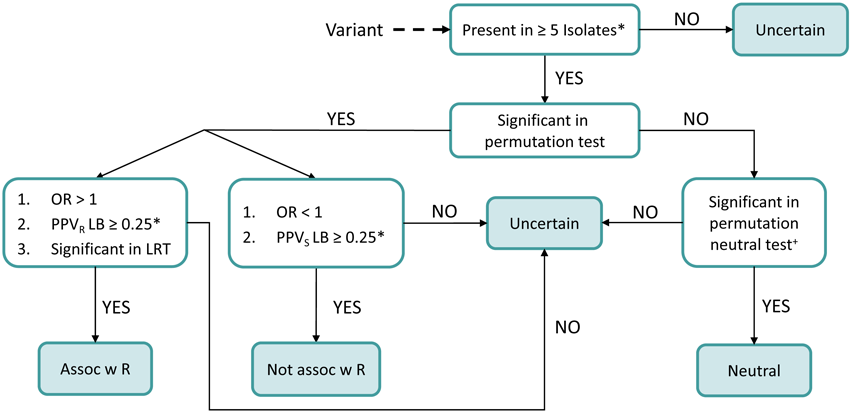
Single-model grading flowchart. Grading steps for a single model. LB = lower bound in a binomial exact confidence interval. *: Relaxed thresholds for *pncA* are the same as in the SOLO algorithm -- Present in ≥ 2 PZA-resistant or susceptible isolates (depending on the sign of the OR) and PPV ≥ 0.5. ^+^: Raw p-values and a cutoff of 0.05 for silent variants in the neutral permutation test. Significance testing in all other cases was performed using FDR- corrected p-values.

To reduce grading of false associations among rare variants, regression-graded variants must be present in at least five isolates. For grading associations with R, we set the FDR q-value cutoff at 0.05 for non-silent variants (including pooled LoF variants), as this constituted the primary analysis, and 0.01 for silent variants to be more conservative in the latter secondary analysis. We additionally required candidate R-associated variants to pass the LRT to mitigate false associations with R, but did not require this for candidate variants not associated with R. For consistency with the SOLO algorithm, we required the lower bound of the PPV to be at least 0.25 for candidate variants associated (OR > 1, PPV_R_) and not associated with R (OR < 1, PPV_S_). PPV_R_ = P(resistant | variant present), PPV_S_ = P(susceptible | variant present), NPV = P(susceptible | variant absent).

The SOLO method set an FDR q-value cutoff of 0.05 for all significance testing, except the Neutral masking algorithm. For consistency with the Neutral masking algorithm of SOLO, we used raw p-values and a cutoff of 0.05 for silent variants in the neutral permutation test.

Like the SOLO algorithm, we integrated associations identified using the WHO dataset (a more select data subset with higher confidence phenotypes) with those from the ALL dataset, which includes the WHO dataset plus additional isolates tested for resistance using phenotypic approaches not endorsed by the WHO. We note that due to differences in the underlying statistical methods, some of the rules for resolving category differences between the two phenotypic groups differ between regression and SOLO^4^.

Importantly, no variants had discrepant associations between the WHO and ALL datasets (*i.e.* graded Assoc w R in one dataset and Not assoc w R or Neutral in the other). 2,388 variants (11%) were not tested in either model due to absence or co-occurrence with low-frequency or missing variant calls, so they were graded Uncertain. Of the 19,201 variants tested in the regression models, 6,710 (35%) were found only in the ALL dataset. Because the WHO dataset is a strict subset of the ALL dataset, no variant was found only in the WHO dataset.

Among the 12,491 variants found in both the WHO and ALL datasets, 12,173 (98%) have the same grading in both datasets. The combined grading across the WHO and ALL datasets reflected the prioritization of resistance calls from the WHO phenotypic testing and associations, but the ability of the ALL dataset to add uncertainty to or upgrade variants that would benefit from higher power or wider sampling **(Supplementary Table 5).** We upgraded variants that were Uncertain in the WHO dataset to Group 2 if the ALL evidence suggested an association with R **(Supplementary Table 5**, row 7). However, in the reverse scenario, the variant was downgraded to Uncertain **(Supplementary Table 5,** row 5) due to the potential for overcalling R-associated variants in the WHO dataset. The SOLO algorithm graded Neutral variants only from the WHO dataset. We therefore prioritized calling Neutrals from the WHO dataset (**Supplementary Table 5,** row 3) by grading them into Group 5, but we additionally upgraded variants that were Neutral in the ALL dataset to Group 4 if they were also present in the WHO dataset **(Supplementary Table 5,** row 4).

Finally, we applied grading rules to the regression output to measure agreement between regression and SOLO + GR, which is the grading of the final published catalog *(World Health Organization,* 2023). To apply grading rules to the regression catalog, we upgraded variants that were graded Uncertain in both regression and SOLO but not Uncertain in SOLO + GR to the SOLO + GR grading.

### G. Resistance prediction and assessing sensitivity and specificity

Both in this work and the published catalogs^3,4^, resistance predictions were made for the same dataset from which the gradings were derived. Gradings were used to predict isolates in the ALL dataset as resistant or susceptible with a presence cutoff of AF > 75%. All isolates were included; no isolates were excluded due to the presence of low-quality or intermediate frequency variants. An isolate containing any variant in Groups 1-2 is predicted resistant, and an isolate lacking all such variants is predicted susceptible. If a pooled LoF variant was graded in Groups 1-2, then all isolates containing any component frameshift, start_lost, stop_gained, or feature_ablation variant in that gene were predicted resistant. We compared metrics between regression, regression + GR, SOLO, and SOLO + GR **(Table 2)** for each drug.

**Table 2.** Counts of gradings for 21,589 unique (drug, variant) pairs, including pooled LoF variants and silent variants, across regression and SOLO, with and without grading rules. The grading group names and interpretations are the same for both SOLO and regression.

| **Group** | **Regression** | **Regression +**<br>**Grading Rules** | **SOLO** | **SOLO +**<br>**Grading Rules** |
| --- | --- | --- | --- | --- |
| 1) Assoc w R | 589 | 589 | 311 | 253 |
| 2) Assoc w R - Interim | 164 | 1,050 | 146 | 1,130 |
| 3) Uncertain | 20,617 | 14,763 | 20,835 | 14,958 |
| 4) Not assoc w R - Interim | 130 | 5,098 | 19 | 4,998 |
| 5) Not assoc w R | 89 | 89 | 278 | 250 |

### H. MIC Models

We built genotypic models on MIC data to investigate and validate associations identified in the binary models. These models did not influence the regression-based grading. For some isolates, multiple MICs measurements were available in different media. We prioritized MICs measured in solid media over liquid media over plate-based assays as detailed in the hierarchy in **Supplementary Table 6** when deduplicating isolates with multiple measured MICs.

All MICs were then normalized to the most common medium for a given drug, which was typically UKMYC due to the large proportion of isolates from the CRyPTIC study^9^. Normalization was done by multiplying the measured MIC by the ratio of the critical concentrations in the most common medium and the medium of the measurement as was previously done^6^. An L2- penalized linear regression model was built regressing the log2-transformed MICs on the same genotypic inputs as for the logistic regression models. The same nested models were fit as for the binary phenotypes, and the regularization parameters were selected by minimizing the root mean squared error. MIC coefficient significance was determined using a permutation test with 1,000 reshufflings and FDR thresholds of 0.05 for non-silent variants and 0.01 for silent variants.

All computation was done in Python 3 using the numpy, pandas, scikit-learn, and statsmodels packages. All code and figures are available at https://github.com/farhat-lab/who-analysis.

## Results

### A. Overview of Regression gradings

After exclusions due to sequencing quality control, the remaining 560 to 48,236 isolates and 31 to 4,086 variants per drug were used to train regression models for 15 drugs (**Supplementary Table 3)**. Phenotypic resistance frequency varied from 2% to 43% across the 15 drugs and differed between the WHO and ALL datasets especially for BDQ (32 percentage points, pp) **(Figure 2a)**. A large fraction of the Bedaquiline (BDQ) pDST data in the WHO dataset came from a single national reference laboratory that received isolates for testing after suspicion of BDQ resistance in source laboratories with limited to no concurrent sampling of BDQ-S isolates^4^. About 80% of all isolates belonged to the Euro-American (L4) and East Asian (L2) genetic lineages (**Figure 2b**).

**Figure 2.**
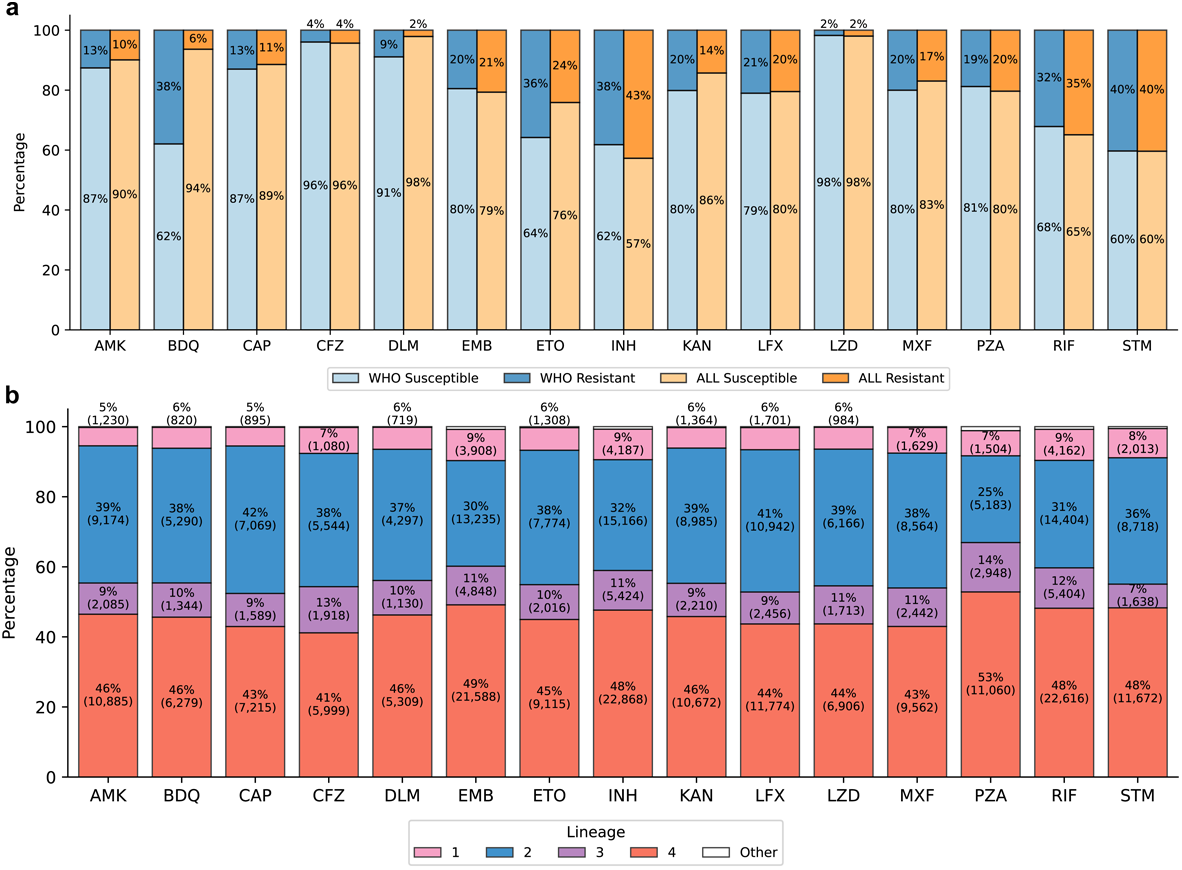
Overview of isolates included in the regression models. a: Percentages of phenotypically resistant and susceptible isolates in the base models for the WHO and ALL datasets, across 15 drugs. **b:** Lineage distribution for isolates in the base model for the ALL dataset only. Other category = *M. bovis* and L5-L7. The percentages for the “Other” category are not shown for readability. Only isolates with a single primary lineage according to the Coll 2014 scheme are shown in panel b. In both panels, isolate counts for each bar are shown in parentheses.

Regression was applied to 21,589 unique (drug, variant) pairs **(Table 2, Supplementary Table 7, Figure 3a).** The resultant catalog was benchmarked against the SOLO data-derived associations without application of additional grading rules (GR). We observe high agreement between the two methods for R-associated variants, with 452/457 (98.9%) of SOLO R-associated variants also detected as such by regression (**Figure 3b).** When GR are applied to regression, it detects 1,380/1,383 (99.8%) of SOLO + GR R-associated variants **(Figure 3c).** Specifically, 804 individual LoF variants are upgraded to Group 2 because the pooled LoF variants for those genes are R-associated, and 5,050 variants (146 non-silent) are upgraded by other GR.

**Figure 3.**
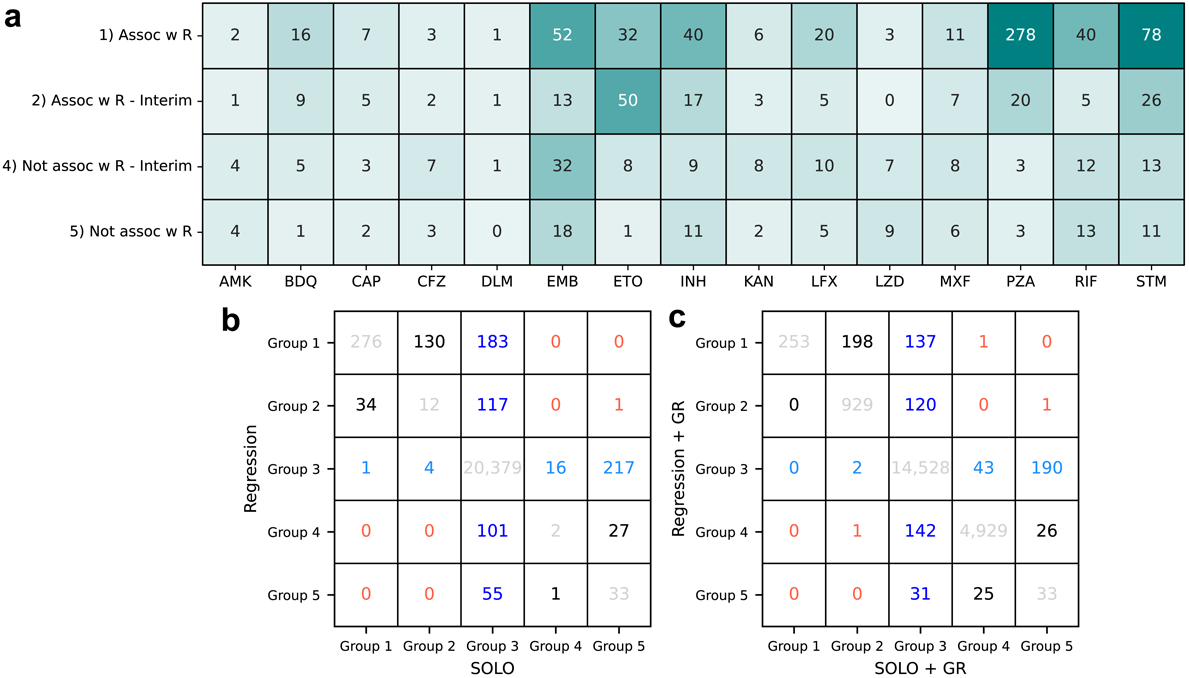
Summary of regression classifications and comparison to SOLO results for 21,589 (drug, variant) pairs. a: Regression variant gradings for 15 drugs, colored by number of variants in each cell. Group 3) Uncertain significance variants are not shown in panel **a**. Grading comparison tables for regression vs. SOLO **(b)** and regression with GR vs. SOLO with GR **(c).** Variant coloring: Dark blue = variants graded Uncertain by SOLO, not Uncertain by regression; light blue = variants graded Uncertain by regression, not Uncertain by SOLO; red = major up-/down-grade discrepancies by regression; gray = group agreement; black = Group 1 or 2 by both regression and SOLO but not perfect agreement.

### B. Sampling and variant co-occurrence affects grading

To understand discrepancies between the gradings, we first examined variants that may have been graded differently due to data or sampling differences between the WHO and ALL datasets. 15 variants were Uncertain in the ALL dataset and Assoc w R in the WHO dataset, resulting in a final grading of Uncertain. In SOLO, 10 of these are graded Uncertain, one Group 1, two Group 2, and two Group 5. The Group 1 variant is Rv0678_p.Met146Thr (BDQ), which is noted to have a potentially inflated PPV due to sampling bias^4^.

Downgrading variants that are Assoc w R in the WHO dataset but Uncertain in the ALL dataset to a final Uncertain grading is exemplified by the variant mmpS5_c.-74G>T (alias Rv0678_c.-11C>A), which has been experimentally associated with BDQ hypersusceptibility in previous work^17,18^. In the binary models, mmpS5_c.-74G>T was measured to have a significant positive association with BDQ resistance in both the WHO and ALL datasets. In the WHO dataset, the PPV LB was also high at 0.77. In the ALL dataset, the PPV LB was less than the Assoc w R threshold (0.25) at 0.19, and this resulted in an uncertain grading. Given the larger size of the ALL dataset, the observed discrepancy raised the possibility of biased sampling in the WHO data, inflating the association with R. The WHO dataset is significantly enriched for BDQ-R with limited to no sampling of BDQ-S isolates from the same communities^4^. This bias is less pronounced in the ALL dataset as evidenced by the lower PPV, but as the WHO dataset is a subset of the ALL dataset, the bias was likely still present **(Figure 2a, Supplementary** Figure 3a). The MIC model, however, reproduced the variant’s known association with S (coef. = −0.043, p-value = 0.008) **(Supplementary Table 10, Supplementary** Figure 3b). Notably, in the BDQ MIC dataset, only 4.4% of isolates are also in the BDQ WHO dataset **(Supplementary Table 2)**. This assessment confirms that biased sampling can skew associations especially for the novel drugs to which resistance remains of limited prevalence globally and is potentially amplified by transmission^19^.

A related methodologically informative observation was that variants causal of resistance can be more diverse and individually rarer than linked polymorphism or mutations associated with hypersusceptibility such as mmpS5_c.-74G>T in the available data. Further, a subset of resistant isolates with the polymorphism may lack potential resistance-causing variants. This results in the regression models assigning a positive effect on drug resistance to the polymorphism or hypersusceptibility variant. Specifically, 11 of 31 BDQ-R isolates with mmpS5_c.-74G>T in the ALL dataset carry one of five Group 1 or 2 BDQ resistance variants graded as such by both SOLO and regression. In addition to mmpS5_c.-74G>T, all 31 isolates share the same three variants in *mmpL5:* Asp767Asn, which is an L2.2.1 marker, Thr794Ile, and Ile948Val, which have been described to be neutral polymorphisms^20^. Only four of 20 isolates without Group 1 or 2 BDQ resistance variants have other variants, all of which were graded Uncertain by SOLO and regression prior to GR. The high proportion of BDQ-R without a potential causal variant suggests the need to look for low-frequency variants below an AF threshold of 0.25 or for variants outside of the evaluated genes for BDQ.

### C. Discrepancies between SOLO and Regression Grading

We next compare gradings across the two methods, identifying no major downward discrepancies, one major upward discrepancy, five minor downward discrepancies, and 233 minor upward discrepancies of regression compared with SOLO (**Figure 3b)**. There are two major discrepancies between regression and SOLO after grading rules are applied **(Figure 3c).**

#### <u>Major downward discrepancies</u> (*i.e.,* Group 4-5 by regression and Group 1-2 by SOLO)

None

#### <u>Major upward discrepancies</u> (*i.e.,* Group 1-2 by regression and Group 4-5 by SOLO)

One variant, rrs_n.514A>C (Capreomycin, CAP). rrs_n.514A>C is a known marker of Streptomycin (STM) resistance^21^; After adding a control for STM resistance in the CAP regression model (by including an additional binary phenotypic STM resistance covariate), rrs_n.514A>C was not significantly associated with CAP resistance, indicating confounding due to a high correlation between CAP and STM resistance as the cause of this discrepancy.

#### <u>Minor downward discrepancies</u> (*i.e.* Uncertain by regression and Group 1-2 by SOLO)

Five variants: Rv0678_p.Met146Thr (BDQ), pncA_p.Ala161fs, pncA_p.Arg154fs, pncA_p.Thr22fs, and pncA_p.Val155Ala (Pyrazinamide, PZA). The first three variants are significant in the permutation test and LRT with OR > 1 but have low PPVs in the ALL dataset. PncA_p.Thr22fs is significant by these tests in the WHO dataset only, and pncA_p.Val155Ala has a permutation p-value of 0.051 in the ALL dataset, putting it slightly over the threshold for significance. As a result, all five variants are graded Uncertain in the ALL dataset and Uncertain overall (**Supplementary Table 5, Methods**).

#### <u>Minor upward discrepancies</u> (*i.e.,* Uncertain by regression and Group 4-5 by SOLO)

There are 233 such variants. Of these, 54 variants are in *rrs* (16S) and were excluded from the regression testing because of higher rates of missing variant calls. This was due to heterozygosity in the sequencing read data and is an issue recognized to arise for ribosomal genes due to homology with other species. Due to rarity (observed in <5 isolates), 26 variants are graded Uncertain by regression, and of these, 16 (all in *rrs* for AMK, CAP, and KAN) have significant FDR≤0.05 in the neutral permutation test. 14 silent variants have OR < 1 with an FDR p-value in the range (0.01, 0.05] but are graded Uncertain because we apply a stricter cut-off of 0.01 for silent variants. The remaining 139 variants meet the SOLO Neutral criterion of PPV_R_ upper bound < 10%^4^ but were not significant in either the regression permutation test or the neutral permutation test.

#### Major discrepancies between regression + GR and SOLO + GR

katG_c.12A>G is graded Group 4 by grading rules because it is a silent variant. It is found in 13 INH-R isolates and 0 INH-S isolates, none of which is a SOLO isolate. Ten isolates have inhA_c.-777C>T, and one has katG_p.Ser315Thr, which are both correctly graded Group 1 by regression. Despite being highly correlated with other INH R-associated variants, katG_c.12A>G is found to be significantly associated with R by regression. This result is similar to that of mmpS5_c.-74G>T and suggests that additional statistical corrections may be required for very high correlations between rare variants. We note that the application of GR corrects this discrepancy for regression.

A single variant is classified as Group 4-5 by regression and Group 2 by SOLO + GR: ethA_p.Pro209fs. This variant only occurs in 26 ETO-S isolates from one lineage (L4.3.4.2), but it is upgraded to Group 2 on grading rules based on the prediction of a frameshift in *ethA* **(Supplementary Table 1)**. Given that ethA_p.Pro209fs is not observed in ETO-R isolates, it either does not have the expected disruption of ethA, or there is an alternative mechanism of activating ETO than the ethA-encoded enzyme in these isolates.

### D. Regression grades 296 (+65%) more R-associated variants than SOLO, superseding the need to use additional grading rules for 78 variants

Regression grades 753 R-associated variants and 219 (160 non-silent) Not Assoc with R variants, compared to SOLO’s 457 and 297 (230 non-silent), respectively and leaves 218 fewer uncertain mutations than SOLO. The use of grading rules is superseded by regression for 78 R-associated variants and 46 Not Assoc with R variants (**Supplementary Table 9**). The 78 variants graded R-associated by regression or by grading rules but not by SOLO are most commonly missed due to a frequency of <5 in isolates without other potential candidate mutations (N=27, 35%) or due to a non-significant association (FDR>0.05 in Fisher’s exact test, N=49, 63%).

The MIC models (**Supplementary Table 10)** were built on largely non-overlapping data with the WHO binary model (up to 15% overlap) **(Supplementary Table 2)** and so allow for partial validation of the binary regression results. Further, as MIC measures resistance more quantitatively, the MIC models can more finely measure the direction and size of effects on resistance, which may more directly link to biological effect^7,22^. We tested 303 (82%) of the 372 variants graded as Assoc of any type by regression and Uncertain by SOLO in the MIC models **(Figure 4)**; the remainder could not be tested due to rarity or the presence of missing or low frequency calls in available isolates with MIC data (MIC dataset set size ranged N=1,007-12,613 across the 15 drugs). 233 variants have a significant regression OR in the WHO dataset, and of these, 79 (34%) are also significantly associated with MIC (73 with resistance, 6 with susceptibility), with all significant associations reproducing the direction of effect measured in the binary WHO models.

**Figure 4.**
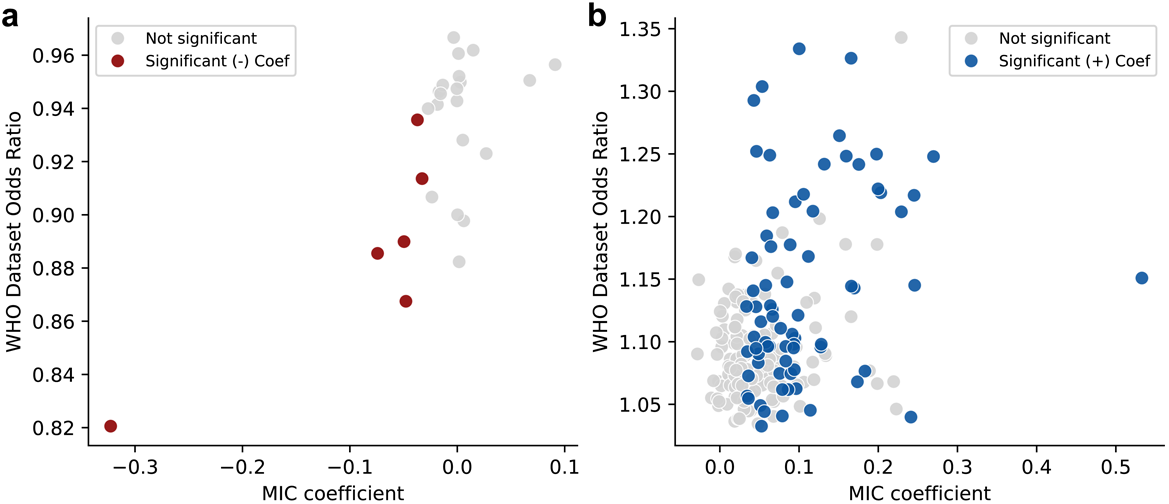
WHO dataset ORs vs. MIC coefficients for 233 variants graded Group 3 by SOLO and Groups 1-2 **(b,** N = 207) or 4-5 (**a,** N = 26) by regression, were tested in MIC models, and have significant OR in the WHO dataset. Neutral variants were excluded. Point color reflects the direction of association in the MIC model and significance at FDR ≤ 0.05 for non-silent variants and FDR ≤ 0.01 for silent variants.

The six drug-variant pairs with significant associations with drug susceptibility in the WHO and MIC datasets (**Figure 4a)** are eis_c.-9T>C (AMK), eis_p.Met1? (loss of start codon) (AMK, KAN), eis_p.His150fs (AMK, KAN), and mmpL5_LoF (BDQ). The literature supports a causal relationship between drug hypersusceptibility and loss-of-function of these two genes. *Eis* is an enzyme that inactivates AMK and KAN, and *mmpL5* is a component of a Bedaquiline efflux pump^23^.

To further probe the newly graded R-associated variants by regression, we inspected RIF R-associated variants by regression that lack a measurable association with MIC and are graded Uncertain by SOLO + GR. There are 5 such variants – F424V, I491L, I491M, I491T, and S493L are missense variants in rpoB outside of the Rifampicin resistance determining region (RRDR). They are predominantly found in RIF-R isolates, but all frequently co-occur with Group 1 variants, most frequently H445N, L430P, and D435Y. Due to the high rates of co-occurrence with known R-associated variants, they have small estimated ORs in the ALL dataset, ranging from 1.06-1.11. They are close to RRDR residues S428 (4.5 Å from F424) and R447-L449 (within 4.6 Å of S493) (PDB ID: 5ZX2, **Supplementary File 1**). For the three variants observed in codon 491, given the diversity of alleles at this site and the known association between I491F and RIF resistance^24^, these associations may be causal.

Some of the newly R-associated variants identified by regression are known INH resistance compensatory mutations. These co-occur with resistance-causing mutations to mitigate their fitness cost relative to wild-type. *katG* detoxifies reactive oxygen species (ROS) and activates the pro-drug Isoniazid. To compensate for the loss of function of katG caused by INH-resistance causing mutations, promoter mutations that increase the expression of *ahpC*, another enzyme with antioxidant functions, are commonly acquired, restoring the bacterium’s resistance to ROS. Regression grades six *ahpC* promoter mutations as R-associated. All are in the region 47-92 base pairs upstream of *ahpC* that is targeted by the Xpert MTB/XDR test^25^, and four have experimental evidence of ahpC overexpression^26-29^ (**Table 3**).

**Table 3.**
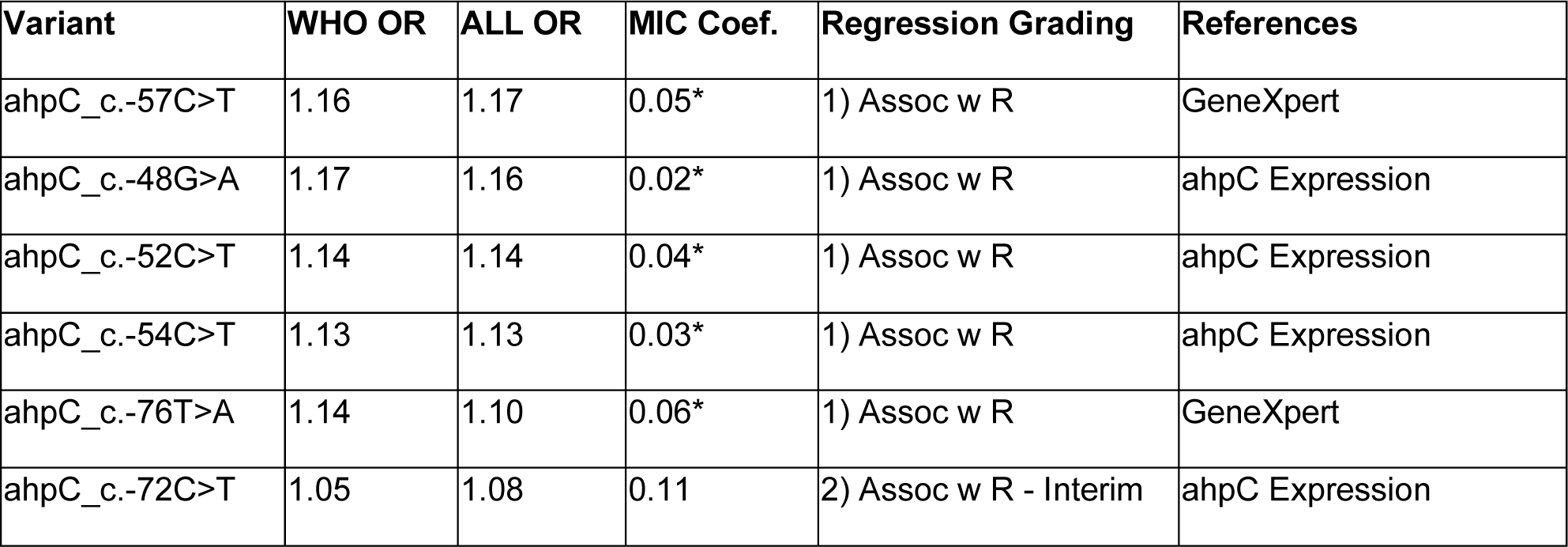
Six isoniazid resistance compensatory mutations derived from ahpC expression data and GeneXpert graded resistance-associated by regression. Variants are ordered by regression grading, then odds ratio in the ALL dataset, in decreasing order. *: ORs or coefficients NOT significant by the corresponding permutation tests. All six variants are graded Uncertain by SOLO and SOLO + GR. The small ORs of these mutations are consistent with indirect effects on INH resistance.

| **Variant** | **WHO OR** | **ALL OR** | **MIC Coef.** | **Regression Grading** | **References** |
| --- | --- | --- | --- | --- | --- |
| ahpC_c.-57C>T | 1.16 | 1.17 | 0.05* | 1) Assoc w R | GeneXpert |
| ahpC_c.-48G>A | 1.17 | 1.16 | 0.02* | 1) Assoc w R | ahpC Expression |
| ahpC_c.-52C>T | 1.14 | 1.14 | 0.04* | 1) Assoc w R | ahpC Expression |
| ahpC_c.-54C>T | 1.13 | 1.13 | 0.03* | 1) Assoc w R | ahpC Expression |
| ahpC_c.-76T>A | 1.14 | 1.10 | 0.06* | 1) Assoc w R | GeneXpert |
| ahpC_c.-72C>T | 1.05 | 1.08 | 0.11 | 2) Assoc w R - Interim | ahpC Expression |

Variants acquired in members of two or more independent lineages are more likely to be causal or compensatory for resistance^30,31^. The majority (177 out of 221) of the variants graded R-associated by regression and Uncertain by SOLO + GR were observed in at least 2 lineages (**Supplementary Table 11, Supplementary** Figure 5). The remaining 44 variants were observed to be lineage restricted, but of these, 18 are rare and found in fewer than 10 isolates, making it less likely to observe acquisition in multiple lineages. (**Supplementary Table 11**).

### E. Sensitivity, specificity, and PPV of the regression-based catalog

We computed sensitivity, specificity and PPV of resistance diagnosis using the catalogs built using SOLO, SOLO + GR, regression, and regression + GR **(Figure 5)**. We expect these performance metrics to be optimistic because they are estimated on the same data from which the catalogs were derived but this derivation is less likely to invalidate the comparison between catalogues. The sensitivity of the regression-derived list is higher for 13/15 drugs than the SOLO-derived list (difference 0.081 to 10.2 percentage points (pp)). SOLO specificity and PPV are higher than regression for 12/15 drugs (specificity difference range: 0.011 to 3.70 pp, PPV difference range: 0.10 to 12.7 pp). For Delamanid, the statistics are the same because both SOLO and regression graded the same two variants as R-associated. The average difference in sensitivity (+3.2 pp) between regression and SOLO is larger than the average differences in specificity (−1.0 pp) and PPV (−1.8 pp), and the F1 scores (harmonic mean of sensitivity and PPV) averaged across the 15 drugs are 69.2% for SOLO, 69.9% for SOLO + GR, and 70.1% for regression.

**Figure 5.**
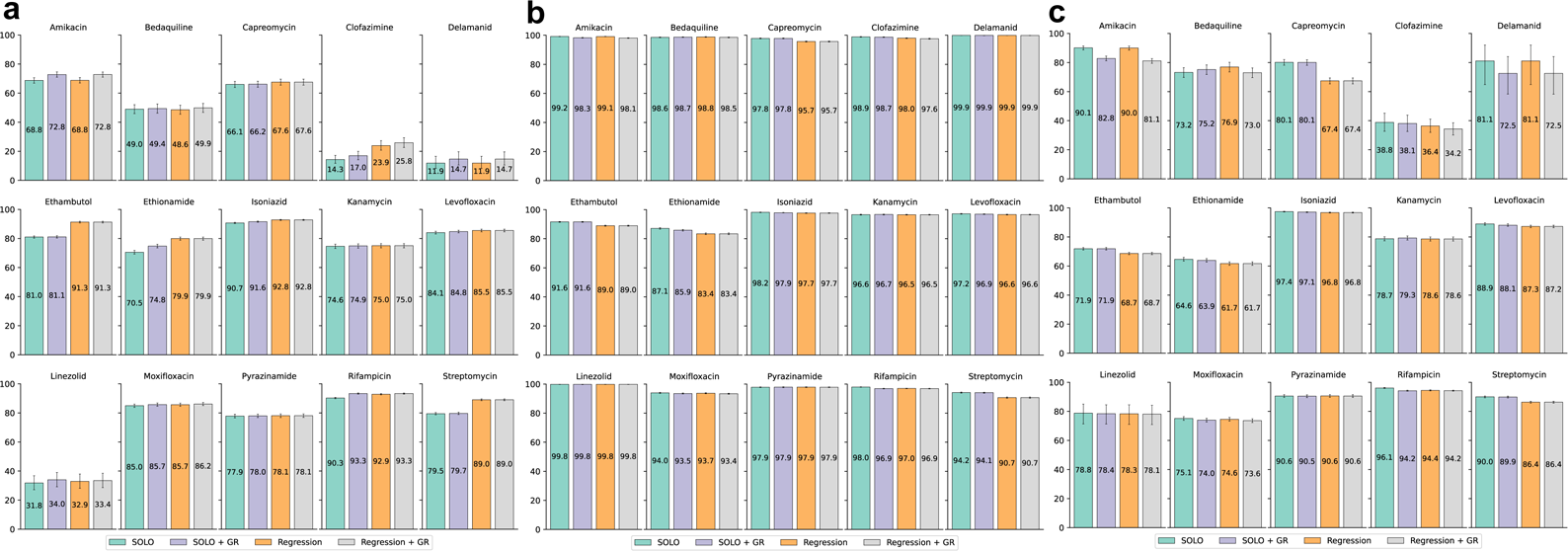
Sensitivity (a), specificity (b), and PPV (c) between SOLO, SOLO + GR, regression, and regression + GR mutation lists. Error bars are 95% exact binomial confidence intervals. Source data are in **Supplementary Table 12.**

The largest gains in sensitivity for regression are for Ethambutol (Δ = +10.2 pp), Clofazimine (+9.6), Streptomycin (+9.6), Ethionamide (+9.4), Rifampicin (+2.6), and Isoniazid (+2.1). For four drugs, regression specificity is more than 1 percentage point lower than SOLO specificity: Ethionamide (Δ = −3.7 pp), Streptomycin (−3.5), Ethambutol (−2.6), and Capreomycin (−2.2). The drop in Capreomycin specificity is due to the rrs_n.514A>C false positive. If this variant is removed from the regression list, the Capreomycin model metrics are the same for both SOLO and regression, and the average F1 score for regression is 70.5% **(Supplementary Table 13).**

When grading rules are applied to the regression results, sensitivity increases for eight drugs, four of which have increases greater than 1 pp. The largest increase is 4 pp for Amikacin due to eis_c.-14C>T and rrs_n.1402C>T, which are upgraded due to WHO approval as resistance markers. Sensitivity increases for Delamanid (+2.8 pp), Clofazimine (+1.9 pp), and Bedaquiline (+1.3 pp), either because of *in vitro* selection evidence (*atpE*, BDQ) or due to LoF mutations (*pepQ,* CFZ) presumed to cause resistance.

The WHO has set minimum target product profiles (TPPs) for genetic drug susceptibility testing – ≥98% specificity for all drugs, and for sensitivity, >95% (RIF), >90% (INH, LFX, MXF), and >80% (BDQ, LZD, CFZ, DLM, AMK, PZA)^32^. For RIF and INH, SOLO specificity is ≥98%, but specificities for SOLO + GR, regression, and regression + GR are <98%. For Clofazimine, the specificity of regression + GR falls below the specificity minimum. For all other drugs, the four methods don’t differ in whether they meet the sensitivity and specificity TPPs.

### F. Lowering AF Threshold to 25% Improves Sensitivities for Bedaquiline, Clofazimine, and Fluoroquinolones

For eight drugs, lowering the AF cutoff to 25% (*i.e.,* variant is considered present if AF > 25%) increased the sensitivity of these mutations for predicting resistant phenotypes by at least 2 pp **(Supplementary** Figure 6**, Supplementary Table 14).** The largest increases are 10.3 pp for Bedaquiline to 58.9% (95% CI, 55.9-61.9%), 4.7 pp for Moxifloxacin to 90.3% (95% CI, 89.4-91.2%), 4.5 pp for Levofloxacin to 90.0% (95% CI, 89.2-90.7%), and 3.8 pp for Clofazimine to 27.8% (95% CI, 24.4-31.3%). The corresponding decreases in specificity are 0.29, 0.36, 0.27, and 0.30 pp, respectively. This confirms the importance of heteroresistance for fluoroquinolones and demonstrates that it is also relevant for Bedaquiline and Clofazimine. These results are consistent with SOLO, for which the largest increase in sensitivity is 10.2 pp for Bedaquiline^4^.

## Discussion

In this work, we demonstrate that logistic regression models can reproduce the results of SOLO, the current WHO catalog resistance-association method in identifying R-associated variants from WGS and resistance phenotype data. Of the 457 R-associated variants graded by SOLO, 453 (99%) are also identified by regression. Regression graded 218 more variants and achieved higher sensitivity on average across 15 drugs than SOLO. Most importantly, regression does not require defining neutral variants before starting grading of mutations. Further, because regression is trained on all high-quality variant data and considers variant co-occurrence, it can reduce the need to use grading rules for variants that do not pass the SOLO algorithm’s inclusion criteria – 124 variants graded by regression required grading rules to be upgraded – or provide evidence against applying grading rules broadly, *e.g.* ethA_p.Pro209fs.

Using the regression pipeline, we identify LoF variants in *eis* and *mmpL5* as associated with aminoglycoside and Bedaquiline hypersusceptibility, respectively, which agrees with recently published work associating genotype with MICs (CRyPTIC Consortium, *Nat Commun*, 2024). We reproduce these associations in MIC models. Overall, we find the MIC models useful as adjunct evidence supporting the gradings derived from the more abundant binary resistance data. The use of MIC and lineage/homoplasy could in the future further reduce the need for grading rules.

The regression grading approach is not without its limitations. Fitting a regression model requires the missing data to be imputed or dropped. Because imputation can introduce bias, we removed isolates containing unfixed variants or variants that could not be called. For the same reasons, variants that were absent from the remaining isolates were also excluded. 2,388 variants (11%) were not fit by the regression models and were automatically graded Uncertain. 64 are putative LoF mutations that were upgraded to Group 2 or 4. Of the remaining 2,324 variants, most (2,270; 98%) are also graded uncertain by SOLO, and 54 (all *rrs:* aminoglycoside pairs) are graded Group 5. Future work in which regression is rerun on subsets of the *rrs* variants and isolates with complete data can address this limitation. Despite the improved ability to grade mutations without the need of additional grading rules, a large proportion of variants (>800 variants) classified in the WHO catalog still require some adjunct evidence to be interpreted.

It is important to note that associations determined by regression do not imply causality. Regression can make false positive associations in the setting of biased sampling, linkage disequilibrium, and high correlation between resistances to different related drugs. Uncovering these limitations informs the application of regression for phenotype association and prediction across a range of systems including in human genetics. That regression can make some false positive relationship may underly its catalogue’s lower specificity and PPV compared with SOLO. However, we also note that the drops in specificity are very small and occur for drugs where phenotypic DST is known to have limited accuracy or reproducibility and where mutations can have intermediate effects on resistance (e.g. ethambutol and ethionamide). Like the approach taken to correct the CAP resistance model for STM resistance (**Results Section B),** future models can correct for resistance to other drugs to reduce these false positives.

To take a completely data-driven approach to classify variants, it is critical to have datasets that are representative of circulating MTBC strains across the spectrum of the resistance phenotype. We observe a considerable range in the percentage of isolates with resistance across the drugs the WHO and ALL datasets with heavy oversampling of resistance compared with its known population prevalence **(Figure 2a, Supplementary** Figure 3a). This was especially the case for bedaquiline. Oversampling is intended to sufficiently sample resistant isolates, but when oversampling is biased, this can result in confounding. In the case of BDQ we observed linkage between neutral polymorphisms indicative of genetic lineage and a diverse set of individually rare resistance variants. Furthermore, several isolates with BDQ resistance are still unexplained by candidate resistance markers **(Results Section B).** We used principal components (PC) to account for lineage effects limiting to the first 50 PCs as there is an expected trade-off between adding additional PCs and statistical power of grading. This PC correction will need to be re-evaluated as more data is available and is expected to be less impactful on power as more representative data becomes available. In the meantime, further analysis of variant lineage distributions and/or phylogenetic inference can help identify the potential false positives arising due to biased sampling and linkage disequilibrium.

In summary, we developed a regression-based pipeline to grade more than 21,000 genetic variants in the MTBC based on their association with resistance to 15 antitubercular drugs. The pipeline is flexible and can be adapted quickly as more data is collected. Although the results we present here are specific to MTBC, this approach provides a model for grading associations between genotype and phenotype for other organisms to use in diagnostic applications.

## Author Contributions

M.R.F. and T.C.R. conceived the study and supervised the research. S.G.K. implemented the code and performed the analyses. The WHO sequencing network contributed raw data. S.L. processed and shared the data. S.G.K. and M.R.F wrote the manuscript. All authors reviewed the manuscript.

## Supporting information

Supplementary Figure and Table Descriptions

## Data Availability

All data produced in the present study are available upon reasonable request to the authors.

https://github.com/farhat-lab/who-analysis/tree/main

## Acknowledgements

We would like to acknowledge Claudio Köser for detailed and helpful feedback on the analysis and writing of the results.

