## Supplementary Figure and Table Descriptions for "Regression for accurate and sensitive grading of mutations diagnostic of antibiotic resistance in *Mycobacterium tuberculosis*"

### Supplementary Tables.

**Supplementary Table 1. Drugs and Tier 1 Genes**. Variants in these genes and the upstream promoter regions were used as input into the regression models. Adapted from Table 21 in the 2nd edition of the WHO mutation catalog *(World Health Organization,* 2023).

**Supplementary Table 2. Model Sizes.** Numbers of isolates and variants dropped due to low-quality or unfixed (within-isolate AF in the range (0.25, 0.75]) variants and final counts used for training each of 135 regression models (9 models per drug). A variant was dropped from the models if, after removing isolates, there were no remaining isolates with that variant present. Pooling combines variants into a single "LoF" variant, reducing the total number of variants. For some models, there was 1 or fewer such LoF variants. Pooling these variants did not affect the model, so the pooled models were not fit. The last two columns show the number and proportion of isolates in the intersection between each model dataset and the WHO dataset. The proportion is equal to the number of overlap isolates divided by the Isolates column.

**Supplementary Table 3. Nested Models.** Nine models were built for each drug using a combination of variant types included in the model (columns) and data subgroups with different phenotype quality criteria and availability (rows). Each model was fit on each set of phenotypes ("WHO dataset," "ALL dataset," and MICs). Red text highlights the model used for grading each specific group of genotypes. Full model sizes, with numbers of isolates and variants, are in Supplementary Table 2. The MIC models were not used for grading; they were only used to add supportive evidence for associations found in the binary drug resistance datasets.

**Supplementary Table 4. Lineages.** Lineage designations for all 52,567 isolates. Fast-lineage-caller was used to assign lineages according to 5 published schemes. The "Lineage" column is the primary lineage based on the Coll scheme, except for eight isolates that could not be classified by this scheme. These eight isolates are *M. canettii* by the Lipworth scheme, and the value in the "Lineage" column is "canettii.". When multiple lineages are called by the Coll scheme, all are reflected in the "Lineage" column.

**Supplementary Table 5. WHO-ALL Grading Integration**. The number of variants across the dataset of 21,589 that meet these criteria are shown in the "# of Variants" column. If a variant is absent from a dataset, it is graded Uncertain. *: Must be present in the WHO dataset; to upgrade variants that are called Neutral only in the ALL dataset to 4) Not assoc w R - Interim, they must not be absent from the WHO dataset. There were no cases when a variant was graded Neutral in one dataset and Assoc w R in the other dataset.

**Supplementary Table 6. MIC Media Hierarchy.** Prioritization of MIC testing media for the regression models. For isolates with MICs tested in multiple media, the media with the highest priority rating was kept. The order is based on ST media prioritization in sections 5·4-5·5 in the 2nd edition of the mutation catalog *(World Health Organization,* 2023).

**Supplementary Table 7. Regression Catalog.** Regression gradings and statistics for 21,589 variants. Data dictionary in Supplementary Table 8. Confidence intervals are 95% exact intervals based on the binomial distribution. For pooled LoF variants that were graded associated with R or not associated with R, all LoF component variants (frameshift, stop_gained, start_lost, and feature_ablation) in those genes were upgraded to Groups 2 or 4 in the “REGRESSION + LOF UPGRADE” column.

**Supplementary Table 8.** **Catalog Column Descriptions.** Descriptions of columns in Supplementary Table 7.

**Supplementary Table 9.** **Mutations for which regression gradings supersede grading rules.** There are 124 variants (78 Group 1-2, 46 Group 4-5) graded by regression but not by SOLO. These are upgraded using grading rules.

**Supplementary Table 10.** **MIC Model Results.** MIC regression results for 9,574 (drug, variant) pairs. All variants tested in the MIC models are given here.

**Supplementary Table 11.** **Lineage Distribution of Regression Upgrades.** Lineage and binary phenotype counts for 219 drug, variant pairs across 12 drugs graded by in Groups 1 and 2 by regression and in Uncertain by SOLO + grading rules.

**Supplementary Table 12.** Source data for **Figure 4.** Binary classification metrics across SOLO = Direct association method, SOLO = SOLO + additional grading rules, and regression. The SOLO statistics are from the published catalog (*World Health Organization,* 2023). All exact 95% confidence interval bounds are included. True positive, true negative, false positive, and false negative counts are also listed.

**Supplementary Table 13.** Binary model results for all 15 drugs with the major discrepancies katG_c.12A>G (INH) and rrs_n.514A>C (CAP) removed. The only differences are for Capreomycin.

**Supplementary Table 14.** Source data for **Figure 8.** Binary classification metrics for AF thresholds of 75% and 25% for regression only. All exact 95% confidence interval bounds are included. True positive, true negative, false positive, and false negative counts are also listed.

### Supplementary Figures.

**Supplementary Figure 1.** Examples of results used to perform the permutation tests and LRT. **a:** Odds ratios for gyrA_p.Asp94Gly (MXF), which is R-associated, in the 1,000 permuted models. **b:** Odds ratios for mshA_p.Asn111Ser (ETO), a neutral masked variant by SOLO, in the 1,000 permuted models. **c:** Example of chi-squared distribution with one degree of freedom (dof) and the area corresponding with the p-value of the LRT. Larger test statistics result in smaller p-values. The dof for the LRT as applied here is one because each time, one variant is removed to test the change in model goodness-of-fit.

**Supplementary Figure 2.** Principal components 1 and 2 **(a)** and 3 and 4 **(b),** colored by MTBC lineage. Principal components show separation between sublineages of L2 (N = 16,801) **(c)** and L1 (N = 4,607) **(d). e-h:** Selected principal components highly correlated with sublineages of L4, showing the diversity of this MTBC lineage. 24,149 isolates have a single L4 sublineage based on the Coll 2014 scheme. For readability, only sublineages that make up at least 2% of this group were plotted, leaving 20,346 isolates across 15 L4 sublineages. Only isolates with a single lineage (but not necessarily a single sublineage) assigned by the Coll *et al.,* 2014 scheme (N = 52,172, 99·25%) are shown here. All lineage classifications are in **Supplementary Table 4.**

**Supplementary Figure 3.** **a:** Percentage of Bedaquiline-resistant isolates (pDST data only), stratified by lineage and phenotypic group. Only isolates with a single primary lineage group assigned are shown here. There are no L5 and *M. bovis* isolates that are BDQ-susceptible. **b:** Distribution of 12,379 available Bedaquiline MICs, stratified by the presence of mmpS5_c.-74G>T. MICs were normalized to the UKMYC5 scale because it is the most common medium in the dataset. The counts of isolates with and without mmpS5_c.-74G>T are normalized separately to better view the much smaller set of isolates that have the variant (N=105). The UKMYC critical concentration for BDQ is 0·25 µg/mL.

**Supplementary Figure 4.** **Composition of the ALL dataset for each drug.** The ALL dataset is composed of WHO-approved pDST results ("WHO dataset"), binarized MICs, and other pDST results not tested using WHO-approved methods. All phenotypic results are shown here, not just those that were included in the regression models. Counts are labeled on the bars, with percentages of each drug's ALL dataset in parentheses. All binarized MICs are part of the ALL dataset. There are no binarized MICs for Capreomycin, Pyrazinamide, or Streptomycin.

**Supplementary Figure 5.** **Lineage and phenotypic distributions for variants graded in Groups 1-2 by regression and Uncertain by SOLO + GR** -- RIF **(a),** INH **(b),** EMB **(c),** LFX **(d),** PZA **(e),** MXF **(f),** ETO **(g),** and STM **(h).** For readability, only the 10 most common variants are shown, and *ahpC* promoter variants covered by the Xpert MTB/XDR test for INH are excluded (Stop TB Partnership, 2024). The lineage distribution is plotted in two columns for each variant: the left column is drug-susceptible isolates (blue), and the right column is drug-resistant isolates (red).

**Supplementary Figure 6.** **Sensitivity (a), specificity (b), and PPV (c) between AF = 75% or 25% as the exclusive cutoff for variant occurrence.** Only results for regression are shown here. Source data are in **Supplementary Table 14**.

### Supplementary Files.

**Supplementary File 1.** **rpoB structure** with variants F424V, I491L, I491M, I491T, and S493L and distances to the rifampicin-resistance determining region.
