## Supplementary figures and images for "Regression for accurate and sensitive grading of mutations diagnostic of antibiotic resistance in *Mycobacterium tuberculosis*"

### Supplementary Figure 1

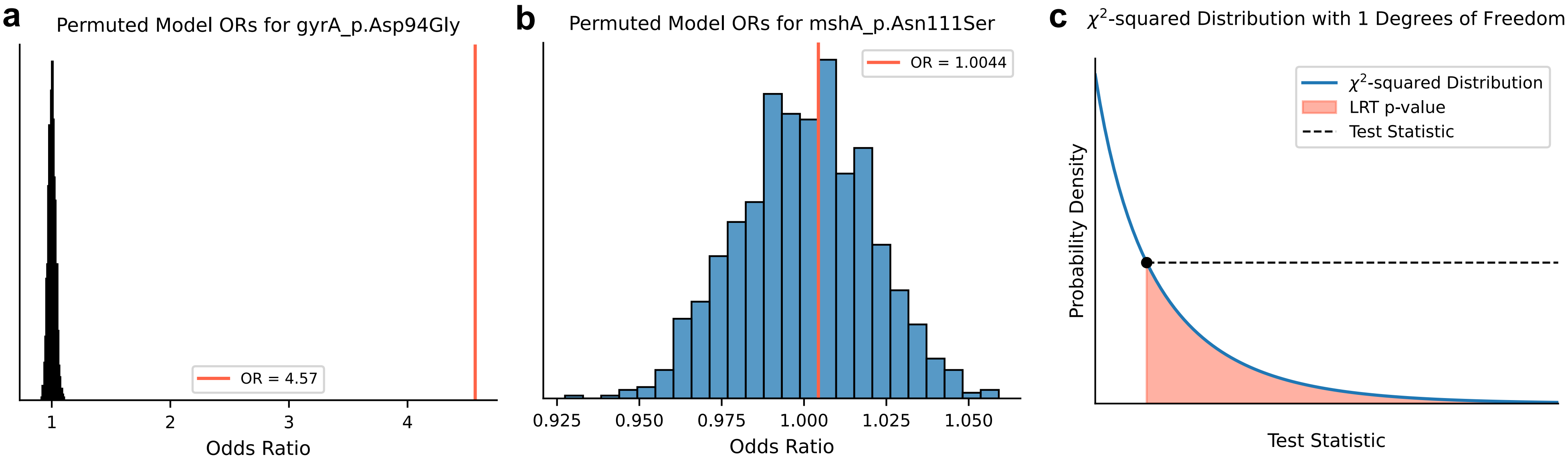

### Supplementary Figure 2

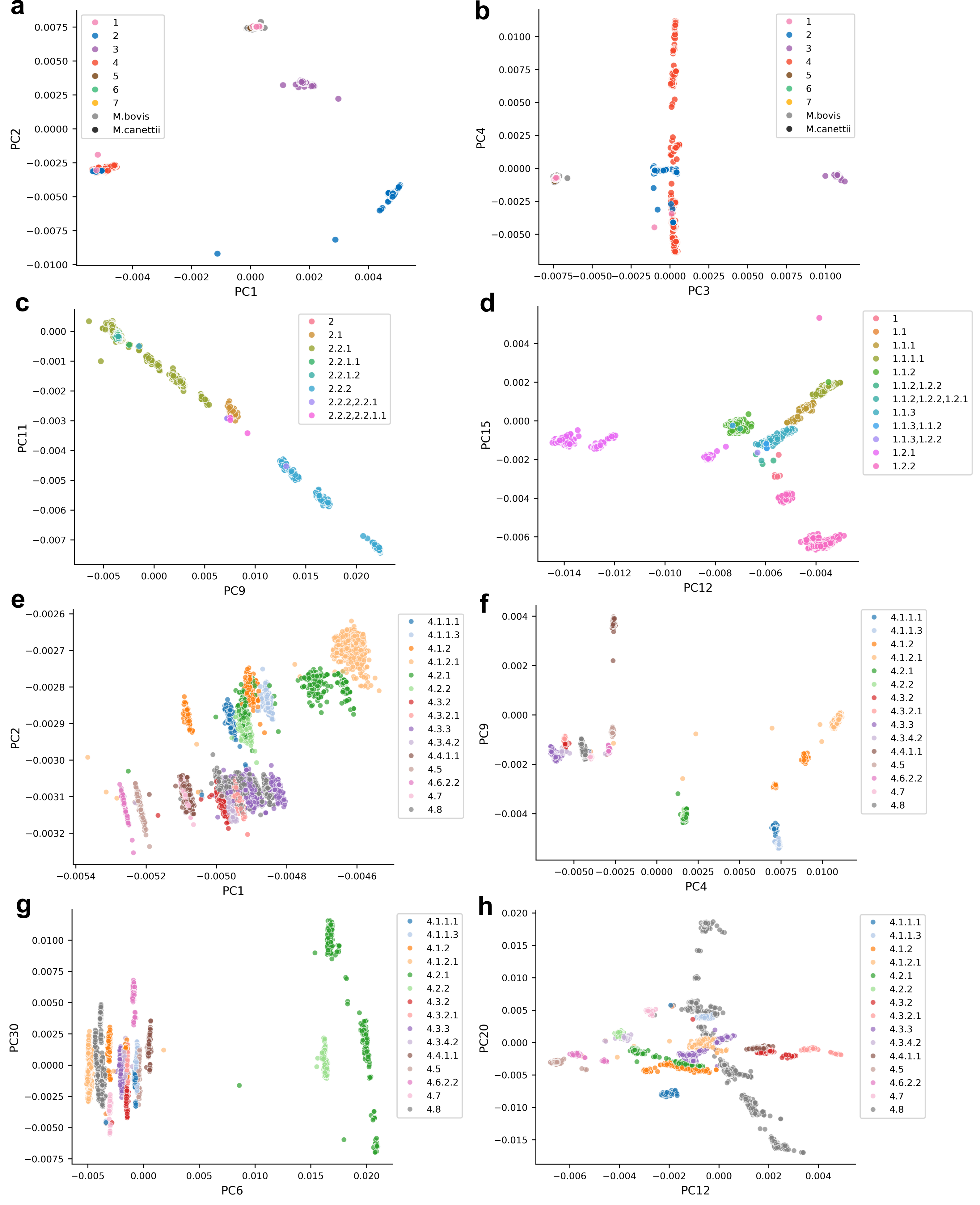

### Supplementary Figure 3

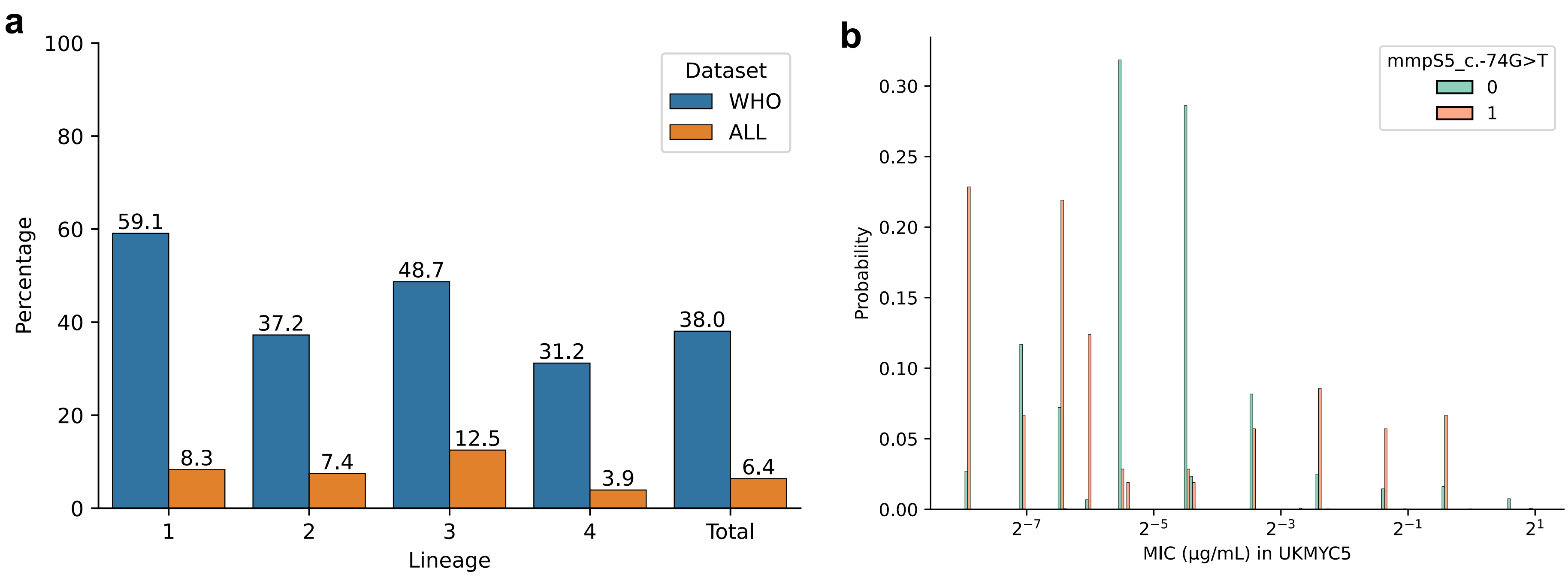

### Supplementary Figure 4

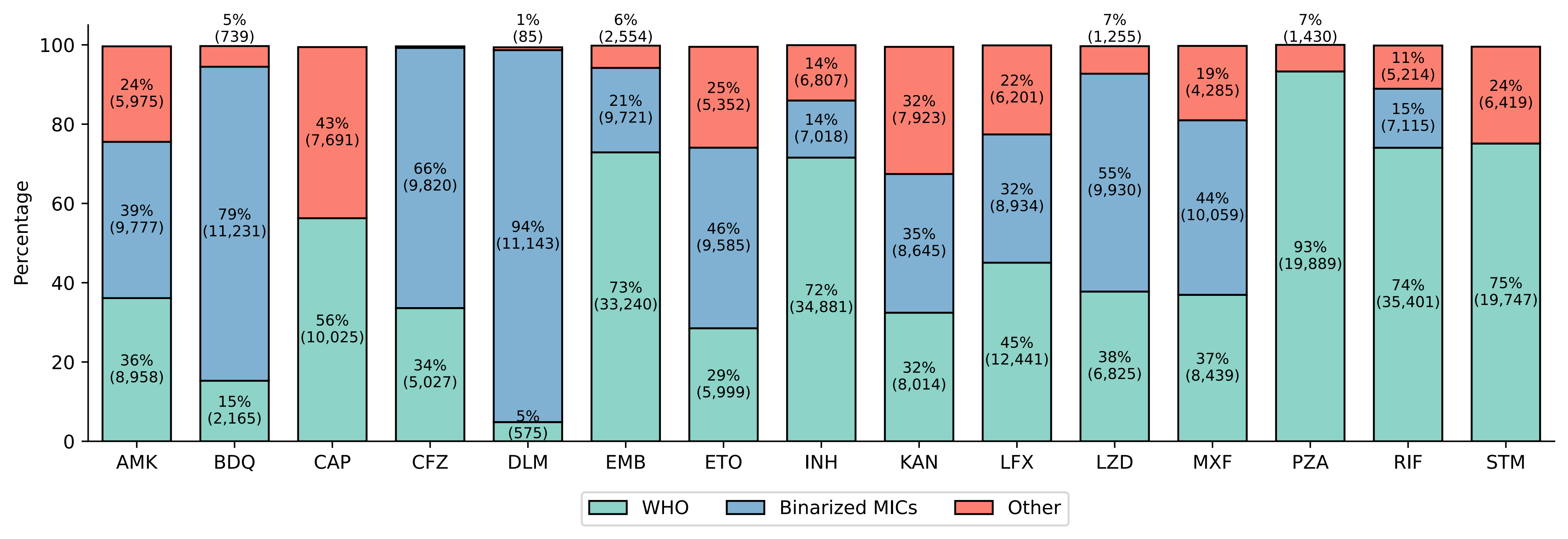

### Supplementary Figure 5

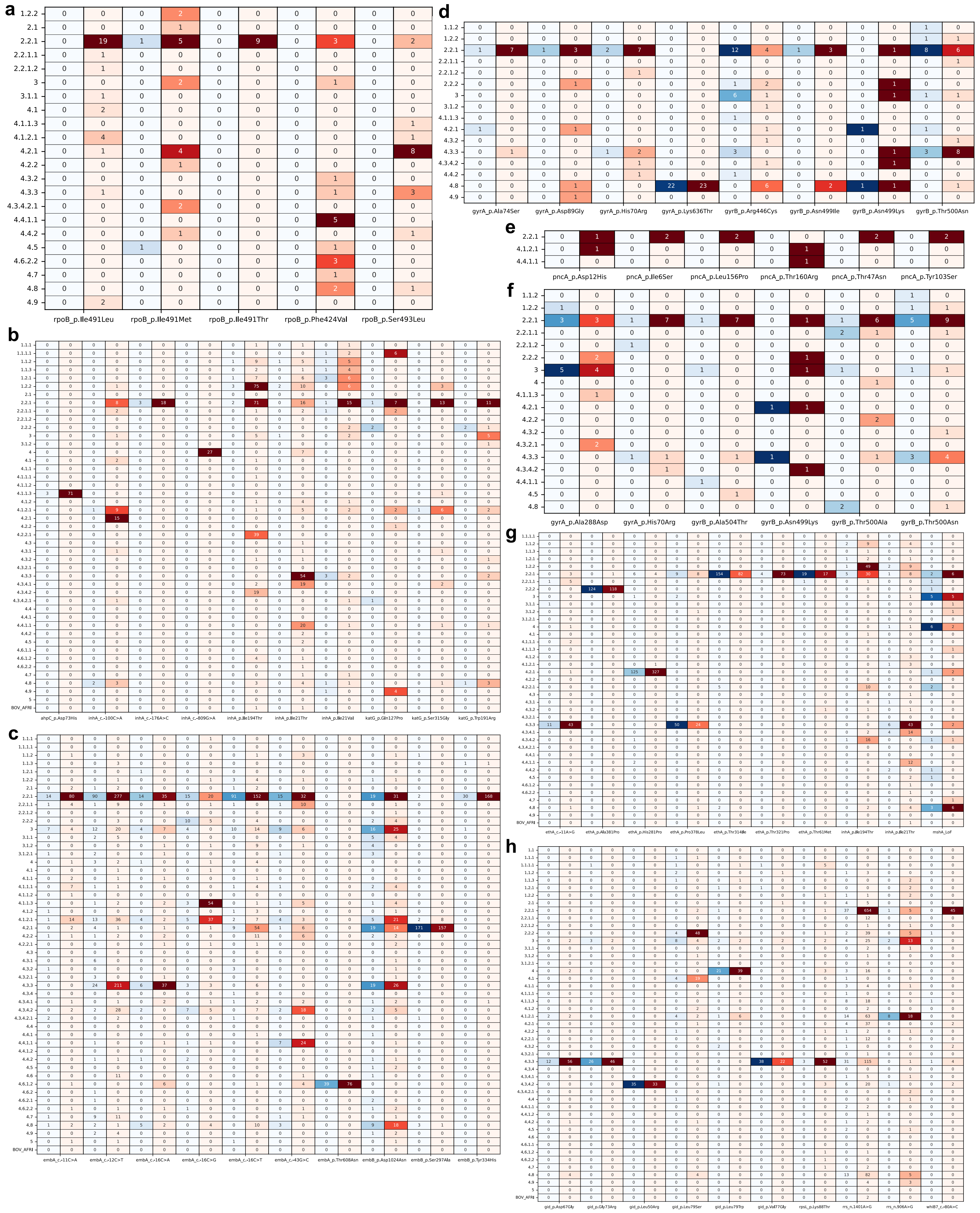

### Supplementary Figure 6

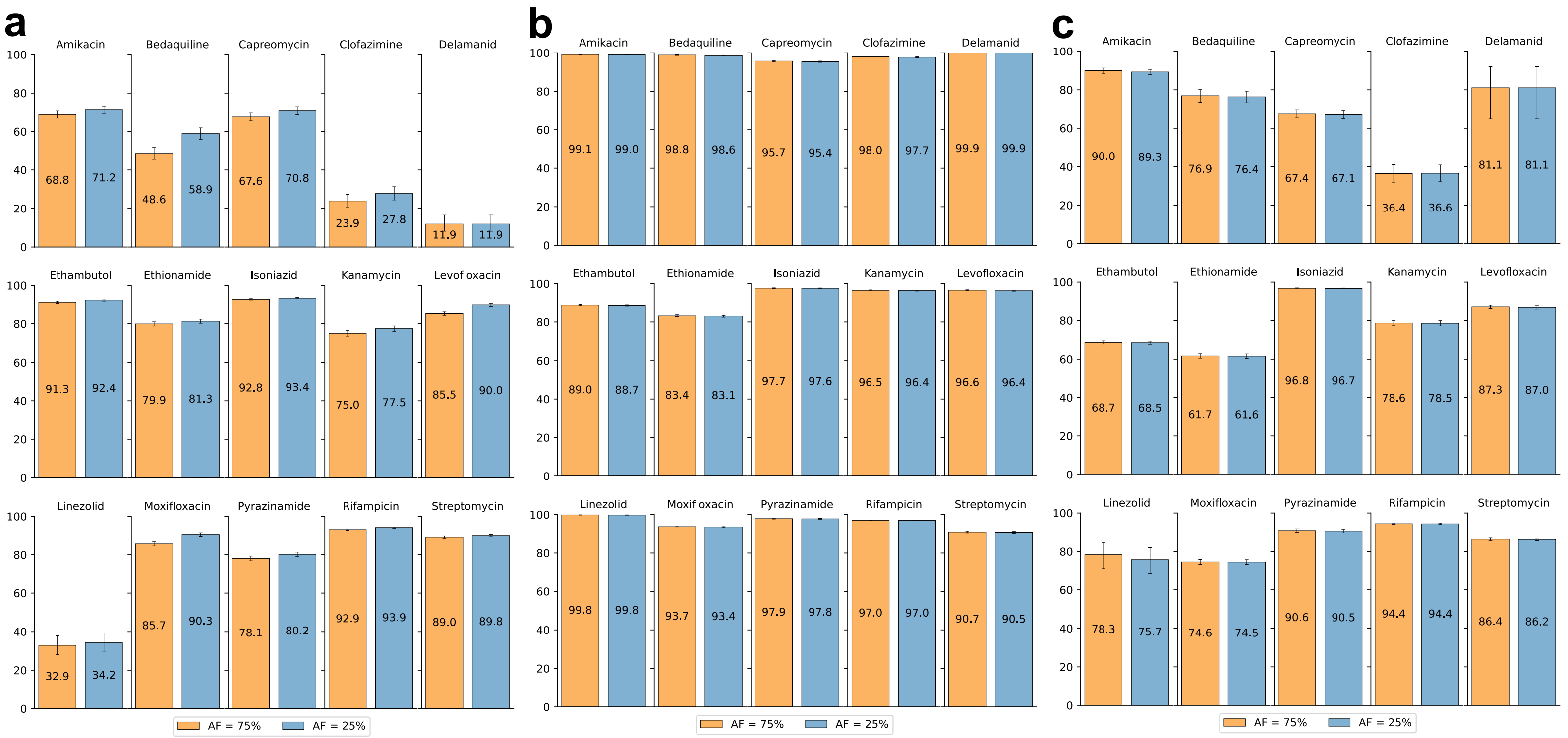
